# PFO Closure in Young-older Patients with Cryptogenic Stroke

**DOI:** 10.1101/2025.10.06.25337460

**Authors:** Dania Mallick, Wanshi Zhong, Ana Torres, Patrik Michel, Alexander Salerno, Muhib A. Khan, Alan T. Davis, Laurel M. Packard, Naser HajAissa, Asad Ahrar, Nadeem Khan, Malgorzata Miller, Nabil Wees, Maulik Kantawala, Tarah A. Moelker, Musa I. Dahu, Mark E. Jacoby, Duane C. Berkompas, William M. Merhi, Min Lou, Jiangyong Min

**Author notes:** Corresponding author: Jiangyong Min, MD, PhD, FAHA, 275 Michigan Street NE, Floor 10, Grand Rapids, MI 49503, USA. These two authors contributed equally to this work (Dania Mallick, Wanshi Zhong).

## Abstract

**Background:** Patent foramen ovale (PFO) closure is recommended for patients younger than 60 years with cryptogenic stroke, including embolic stroke of undetermined source (ESUS). However, the role of PFO closure in patients older than 60 years remains uncertain. We aim to investigate the benefits of PFO closure in young-older patients (aged between 60 and 70 years) with cryptogenic stroke.

**Methods:** Patients older than 60 years with cryptogenic stroke from large tertiary hospitals who received a PFO closure after a multidisciplinary brain-heart case conference, compared to an age-matched cohort of patients with cryptogenic stroke. Demographcis, vascular risk factors, and risk of paradoxical embolism (RoPE) score were collected for comparison between two groups. The primary outcome included recurrent ischemic stroke that occurred up to 5 years following PFO closure between the two groups. The safety outcome was the occurrence of any complications after PFO closure, including new-onset atrial fibrillation (AF).

**Results:** A total of 102 patients with cryptogenic stroke were included (mean age 66±4), 56 male (55%), who had a PFO closure. Procedure-related complications, including new onset transient AF, were low (1%). There was no statistical difference between two groups in patients demographic and clinical characteristics, but the RoPE score was significantly higher in the PFO closure group than in the control group. After a median follow-up of 54 months, patients in the PFO closure group demonstrated a relatively low incidence of recurrent ischemic stroke (3.9%, 4/102), compared to an age-matched control cohort with a median follow-up of 51 months, which had a higher recurrence rate (8.2%, 8/97).

**Conclusions:** In patients with cryptogenic stroke who are older than 60 years, PFO closure demonstrated safety profiles with a trend of reducing recurrent ischemic stroke compared to their age-matched counterparts. Prospective randomized trials are warranted in this young-older population with cryptogenic stroke and PFO.

## Introduction

Patent foramen ovale (PFO) closure has emerged as a key intervention in selecting patients with cryptogenic stroke, particularly younger individuals (age < 60 years) with cryptogenic stroke including embolic stroke of undetermined source (ESUS) (1, 2, 3, 4,5). However, its role in older patients (age >60 years), particularly in young-older (age between 60 and 70 years), remains a subject of debate due to the increasing prevalence of alternative stroke mechanisms, such as atrial fibrillation (AF) and atherosclerosis, in this population. Despite these concerns, recent limited evidence suggests that PFO may still play a pathogenic role in cryptogenic stroke among young-older patients with cryptogenic stroke, warranting careful patient selection for PFO closure.

Several randomized notable clinical trials (RCTs), including percutaneous closure of PFO using the Amplatzer PFO occluder with medical treatment in patients with cryptogenic embolism (PC) (1), PFO closure to established current standard of care treatment (RESPECT) (2), PFO closure or anticoagulants versus antiplatelet therapy to prevent stroke recurrence (CLOSE) (3), Gore septal occluder device for PFO closure in stroke patients (REDUCE) (4), and device closure versus medical therapy for cryptogenic stroke patients with high-risk PFO (DEFENSE-PFO) (5), have established the benefit of PFO closure in younger patients (ages 18–60) with cryptogenic stroke, demonstrating a significant reduction in recurrent stroke compared to medical therapy alone. However, these trials excluded older patients, leading to uncertainty regarding the efficacy and safety of PFO closure in those aged over 60.

Emerging observational studies and subgroup analyses suggest that well-selected older adults with cryptogenic stroke and high-risk PFO morphology may still derive benefit from closure, especially when other stroke mechanisms are ruled out (6), but convincing data remain limited. Nonetheless, patient selection is crucial in these relatively old patients, given the higher prevalence of age-related comorbidities, including occult subclinical AF, which may contribute to stroke recurrence.

Understanding the role of PFO in older adults is critical to refining treatment strategies and optimizing stroke prevention in this special population of young-older adults. This observational cohort study aimed to evaluate the benefit of PFO closure in young-older cryptogenic stroke patients aged between 60 and 70 years, compared to an age-matched control group receiving the best medical management for preventing recurrent stroke. In addition, the safety outcome was evaluated to balance the risk versus the benefit.

## Methods

This study was an investigator-initiated, observational, multicenter collaboration. We identified patients from our daily practice and stroke registry who were consecutively admitted to large tertiary hospitals with the diagnosis of acute ischemic stroke (AIS) from January 1, 2017, to December 31, 2020. The study proposal was approved by the institutional review board (IRB). Patients were eligible if they were 60 years or older with a verified diagnosis of cryptogenic by magnetic resonance imaging (MRI) following an extensive stroke workup and multidisciplinary approach (7). PFO was diagnosed by transthoracic and transesophageal echocardiography with agitated saline. Patients were excluded if their stroke etiology was determined as a result of small-vessel disease or intracranial artery stenosis, large artery atherosclerosis, stroke of other determined etiology, or other cardioembolism but not PFO. Patients who take anticoagulation in the setting of a hypercoagulable disorder for various reasons were excluded. Patients were also excluded if they had uncontrolled diabetes mellitus (DM), uncontrolled hypertension (HTN), autoimmune disease, or a recent history of alcohol or drug abuse. A minimal 28-day cardiac monitoring was applied to rule out asymptomatic subclinical AF or atrial flutter. If a patient had two of these common AF risk factors (HTN, DM, obstructive sleep apnea [OSA], obesity, congestive heart failure, coronary artery disease, and alcohol abuse), insertable prolonged cardiac monitoring was pursued for a minimum of 6 months before PFO closure. Transient ischemic attack (TIA) was excluded from this study cohort to minimize patient selection bias. Patients’ demographic characteristics, vascular risk factors, initial National Institutes of Health Stroke Scale (NIHSS), and RoPE score were collected for comparison. Amplatzer PFO Occluder (Abbott Cardiovascular, Plymouth, MN) was used in all closure patients. Dual antiplatelet therapy (DAPT) was initiated after PFO closure for 90 days, followed by long-term single antiplatelet therapy (SAPT) indefinitely, after a repeat echocardiography 90 days following PFO closure revealed a well-seated device with no evidence of residual shunt. The primary outcome included recurrent ischemic stroke that occurred up to 5 years following PFO closure. The safety outcome was the occurrence of any complications after PFO closure, including post-procedural new onset AF.

Statistical analysis: Quantitative data are expressed as the mean+SD, while ranked data (Initial NIHSS, Baseline mRS) are expressed as the median (minimum, maximum). Nominal data are expressed as a percentage. Comparisons between groups for quantitative variables were performed using the two-tailed, unpaired t-test, while ranked data were analyzed using the Mann-Whitney test. Nominal variables were evaluated using the chi-square test. Determination of the differences between the PFO group and the medical treatment group for the probability of freedom from recurrent stroke was performed using the log-rank test, while the graphical representation was performed using the Kaplan-Meier method. The hazard ratio was evaluated using the Cox proportional hazards model. For this analysis, the dependent variable was the time to recurrent stroke, while the independent variable was treatment (medical treatment or patent foramen ovale closure). Significance was assessed at p<0.05. All analyses were performed using Stata v.17.0 (StataCorp, College Station, TX).

## Results

A total of 102 patients (age >60 years old) with acute cryptogenic stroke were included in the study and compared to 97 age-matched cryptogenic stroke patients who received the best medical management. There was no statistical difference between the group of patients with PFO closure and the control group, except for race and the RoPE score (Table 1). The PFO closure group consisted predominantly of Caucasian patients, whereas the control group was composed mainly of non-Caucasian patients (Table 1). The median RoPE score was 5 (4.6±1.0) in the PFO closure group, which is significantly higher than the control group’s RoPE score of 4 (3.8±1.0) (Table 1).

**Table 1.**
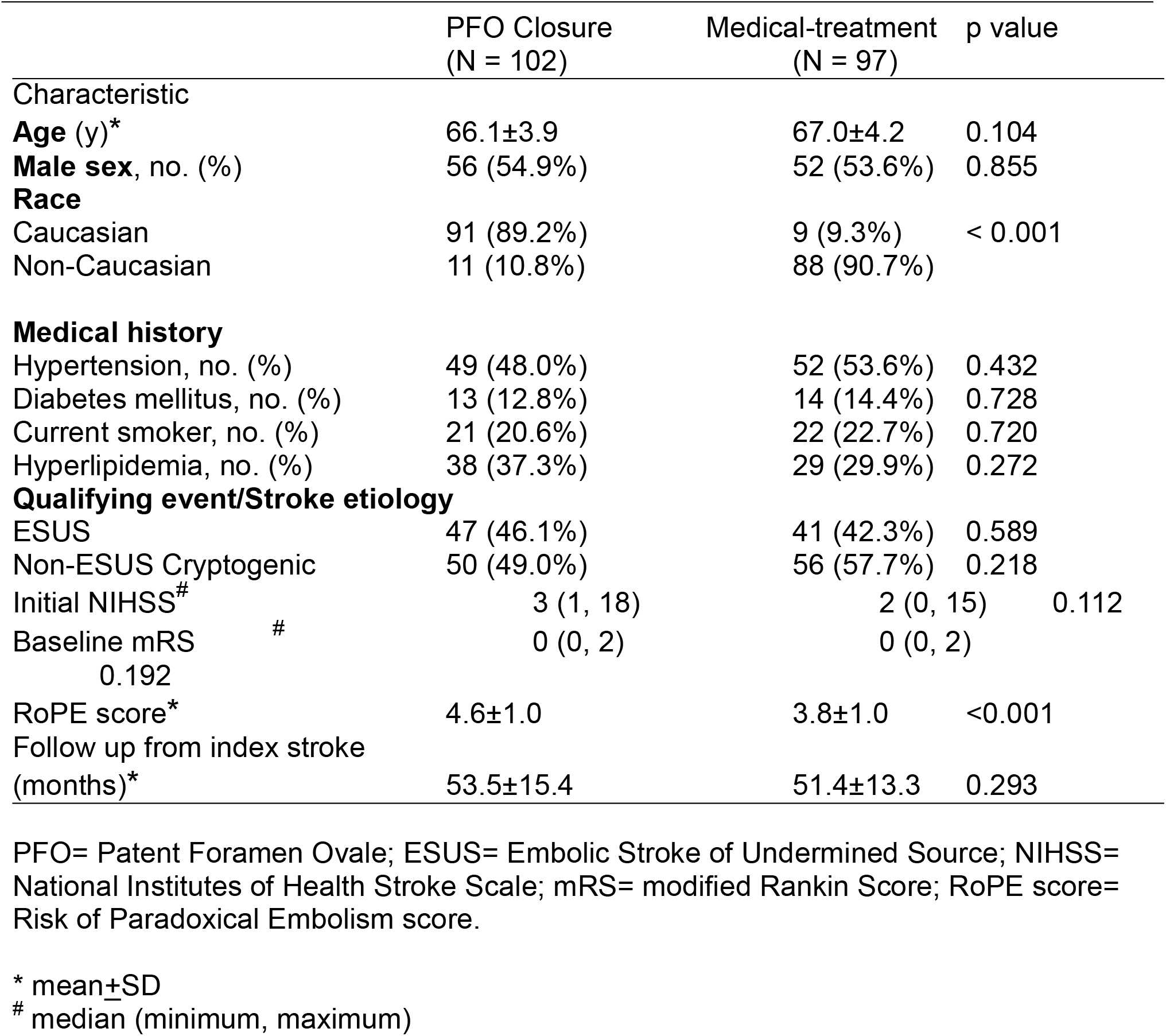
Baseline Characteristics of Patients.

Efficacy and safety outcomes were summarized in Table 2. Implantation and retention of the PFO occluder were successful in all 102 patients (100%) in the PFO closure group. No post-procedural complications occurred in patients who received PFO closure, except one patient developed transient AF after the procedure, which lasted for a few minutes, with no recurrent AF with the following one-year cardiac monitoring. No cerebral hemorrhage or death occurred after the method of PFO closure and subsequent follow-up in the two groups. Recurrent ischemic stroke occurred in 4 patients (3.9%) in the PFO closure group during the median follow-up of 54 months, and in 8 patients (8.2%) during the medium follow-up of 51 months in the medical management group. A notable trend of recurrent stroke reduction is reflected in the Kaplan-Meier curves for the PFO closure group compared to the medical treatment group (hazard ratio for recurrent stroke, 0.43; 95% confidence interval [CI], 0.13 to 1.42; P = 0.152) (Figure 1), although statistical significance was not achieved.

**Table 2.** Efficacy and Safety Outcomes.

| **Primary efficacy** | **PFO Closure** | **Medical Treatment** |
| --- | --- | --- |
|  | **(N = 102)** | **(N = 97)** |
| Recurrent stroke<br>(TOAST classification) |  |  |
| SVD | 2 | 2 |
| Large vessel | 1 | 2 |
| Cryptogenic (ESUS, %) | 1 (1%) | 4 (4%) |
| Cardioembolism | 0 | 0 |
| Stroke of other determined cause | 0 | 0 |
| Fetal stroke | 0 | 0 |
| Systemic embolism | 0 | 0 |
| **Safety outcomes** |  |  |
| Cerebral hemorrhage | 0 | 0 |
| Death from any cause | 0 | 0 |
| **Success of device implantation** |  |  |
| No. (%) | 102 (100%) | N/A |
| Success of PFO closure |  |  |
| No. (%) | 102 (100%) | N/A |

TOAST= the Trial of ORG 10172 in Acute Stroke Treatment classification; SVD= small vessel disease. Due to small recurrent events, running a standard hazard ratio test is not feasible.

**Figure 1.**
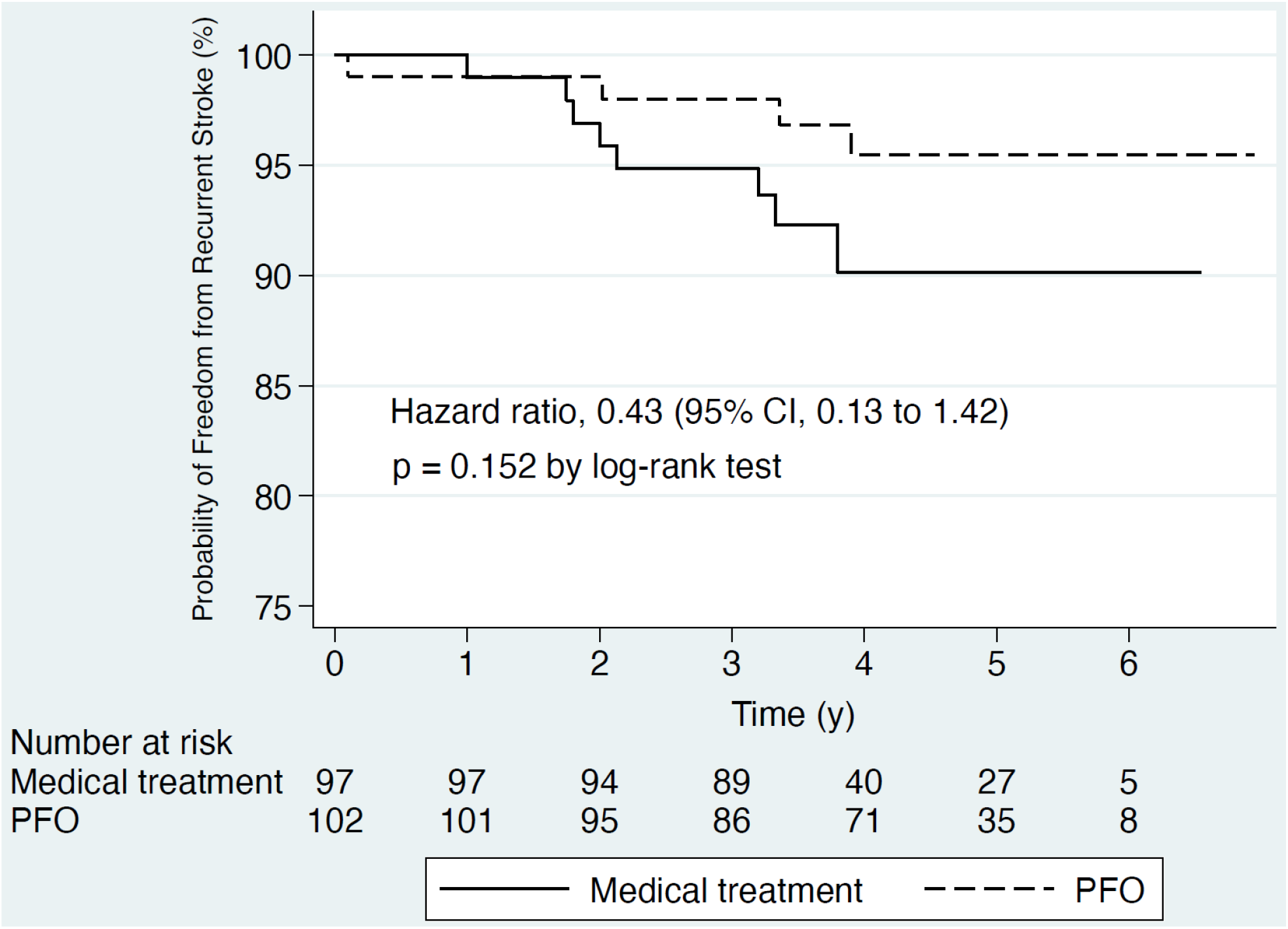
Kaplan–Meier cumulative estimates of recurrent stroke probability were compared between the PFO closure group and the medical treatment group. The analysis was conducted in the intention-to-treat cohort, encompassing all study participants. Although the difference did not reach statistical significance based on the log-rank test, a trend toward a lower incidence of recurrent stroke was observed in the PFO closure group compared to the medical treatment group.

Based on the TOAST stroke classification (8) and evaluated by stroke neurologists, two patients with recurrent stroke in each group were determined to have small vessel disease. One patient in the PFO and two patients with recurrent stroke in the control group had their stroke etiology deemed as large vessel disease. In addition, one in the PFO closure group and four in the control group had cryptogenic stroke determined by stroke neurologists. No fetal stroke or systemic embolism occurred in this study cohort.

## Discussion

Our multicenter observational study demonstrated a trend of reduction of recurrent ischemic stroke with no difference in a composite of systemic embolism, cerebral hemorrhage, or death in patients with PFO closure compared to an age-matched control group in the young-older population (60-70 years old) with cryptogenic stroke or ESUS at a median of 53 months post-procedure follow-up. No procedural complications occurred in our study cohort except one patient developed minutes of transient asymptomatic AF after the PFO closure with no recurrence, and all patients received successful PFO closure.

Major PFO closure trials (1, 2, 3, 4, 5) that established the benefit of PFO closure largely excluded patients older than 60 years. The AHA/ASA (9) and the European Stroke Organization (ESO) (10) endorse PFO closure for individuals between 18 and 60 years old. However, no definitive randomized clinical trials (RCTs) have been conducted in patients aged 60 or older with cryptogenic stroke. Interestingly, a subgroup analysis from the DEFENSE-PFO trial (11), which included patients aged 18 to 80 years, evaluated the outcomes of patients 60 years or older who received PFO closure compared to those who received medical management. All recurrent ischemic stroke or TIA occurred in patients with medical therapy, and none of them happened in patients who had PFO closure. Nonrandomized observation studies (12,13) showed that PFO closure was safe and associated with a relatively low incidence of recurrent ischemic events in older patients compared to historical cohorts of younger patients with cryptogenic stroke who received medical treatment. Authors (12,13) suggest that PFO closure may benefit carefully selected older patients with cryptogenic stroke. A statement from the Roundtable of Academia and Industry for Stroke Prevention (RAISE) (14) emphasized there is a knowledge gap in cryptogenic stroke patients older than 60 years with PFO. Dr. Sposato and colleagues (14) stated that clinical trials are needed to assess the efficacy and safety of PFO closure compared to medical management in cryptogenic stroke patients aged above 60 with PFO. A recent meta-analysis (15) demonstrated similar benefits from PFO closure in recurrent stroke prevention for both older (≥60 years) and younger (<60 years) cryptogenic patients with PFO.

Our study is the first observational study providing the long-term primary outcome of recurrent stroke and the safety outcome of peri-procedural complications compared to the age-matched control group who received medical treatment. With a careful and well-defined evaluation process by the multidisciplinary approach, including stroke neurologists and structural cardiologists, we found that cryptogenic stroke patients with PFO closure are more effective than medical management for preventing recurrent stroke. RoPE score is higher in the PFO closure group than in patients who received medical treatment (with a mean of 5 versus 4, respectively). Of note (Table 2), only one (1%) recurrent cryptogenic stroke occurred in the PFO closure group versus four (4%) in the group of medical treatment. Due to the very low incidence of recurrent cryptogenic stroke, statistical analysis was unable to be conducted. In contrast to the reported high incidence of cardiovascular events and mortality rate among older patients by Wintzer-Wehekind and colleagues (12), no procedure or device-related complications occurred in our study cohort. This excellent safety profile could be attributed to advanced devices and the expertise of skilled structural cardiologists.

The present study has several limitations. First, the sample size is relatively small, so it may not accurately reflect the circumstances in real-world practice. A trend of low recurrent stroke in the group of PFO closure compared to medical treatment was noted in this study. However, the result could be different with an increasing sample size. Second, morphometric features of PFO, socioeconomic status, sex differences, and the underserved patient population were not addressed in this study. Third, patients with PFO closure were predominantly Caucasian, whereas the control group of patients was mainly non-Caucasian. Therefore, we should carefully extend the current result to different populations.

In conclusion, PFO closure could remain effective in preventing stroke among young-older patients (age 60–70 years old) with careful patient selection via a brain-heart board multidisciplinary approach. Randomized trials involving more patients with age-matched control subjects are surely required to validate our observational findings.

## Data Availability

The data that support the findings of this study are available on request.

## Data Availbility Statement

The data that support the findings of this study are available on request.

